# Detection of Anti-H5 Antibodies in People with Exposure to Wild Birds in Northern Canada

**DOI:** 10.64898/2026.05.24.26353994

**Authors:** Hannah L. Wallace, Morgan Hiebert, Mikayla Hunter, Megan Halbrook, Ryan J. Harrigan, Isaac I. Bogoch, Anne W. Rimoin, Souradet Shaw, Linda Larcombe, Pamela H. Orr, Jason Kindrachuk

**Author notes:** Corresponding Authors: Jason Kindrachuk, Pamela H. Orr, Linda Larcombe. Authors contributed equally to this article (Hannah L. Wallace, Morgan Hiebert). Supervised equally (Isaac I. Bogoch, Anne W. Rimoin, Souradet Shaw, Linda Larcombe, Pamela H. Orr, Jason Kindrachuk).

## Abstract

Using a commercially available H5 serology assay, we identified a 7.4% (*n*=5/68) anti-H5 seroreactivity rate among hunters in Northern Canada. All participants reported close contact with wild birds.

## Introduction

Highly pathogenic avian influenza (HPAI) virus H5N1 circulates in wild bird populations, causing sporadic outbreaks in poultry operations, non-human mammals, and occasionally spilling over into human populations. Multiple introductions of HPAI clade 2.3.4.4b into North America have occurred since late 2021, and have caused substantial outbreaks in wild birds, non-human mammals, and poultry (1,2). This clade has undergone broad global expansion since 2021, infecting a wide array of species, with detection on every continent except for Oceania. In 2024, HPAI H5N1 was detected in dairy cattle for the first time in the United States, characterized by sustained cattle-to-cattle transmission and several dairy-associated human cases (3–6).

Following the introduction of H5N1 avian influenza clade 2.3.4.b into North America in 2021 (7), infections among mammals, including humans, have increased during the wide-scale spread of this virus among birds (8,9). Given this increase, it is suspected that individuals who interact with wild birds may be at elevated risk for infection. However, the true incidence of human H5N1 infections is unknown (10). The aim of the current study was to assess the prevalence of anti-H5 antibodies among populations at increased risk for infection in Northern Canada.

### The Study

In August 2025 (three months after the 2025 spring goose hunting season), 68 dried blood spot (DBS) samples with matched questionnaires (Appendix 1) were collected from a northern community in Manitoba, Canada. Recruitment was focused on those > 18 years old who were involved in the hunting of birds and/or preparing them (plucking, butchering, food preparation) for consumption. Some of the participants were also involved in hunting, trapping and handling of mammals (**Table 1**). Participants were recruited through social media posts and community posters. Participation was voluntary, and included both local community members as well as individuals who were visiting the community for the upcoming fall goose hunting season. Participants were compensated for their involvement in the study. This study was approved by the University of Manitoba Health Research Ethics Board under study HS26941 (H2025-151).

**Table 1.**
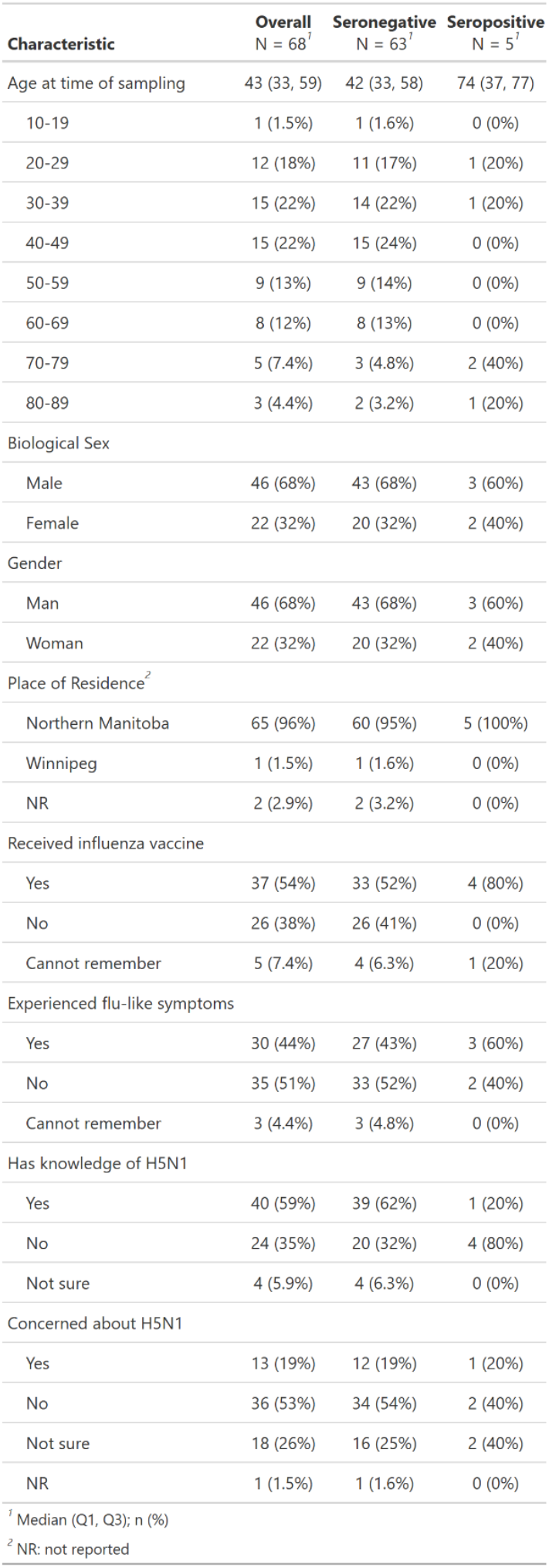
Demographic data and knowledge of HPAI H5N1 for all participants.

| Characteristic | Overall<br>N = 68<sup>1</sup> | Seronegative<br>N = 63<sup>1</sup> | Seropositive<br>N = 5<sup>1</sup> |
| --- | --- | --- | --- |
| Age at time of sampling | 43 (33, 59) | 42 (33, 58) | 74 (37, 77) |
| 10-19 | 1 (1.5%) | 1 (1.6%) | 0 (0%) |
| 20-29 | 12 (18%) | 11 (17%) | 1 (20%) |
| 30-39 | 15 (22%) | 14 (22%) | 1 (20%) |
| 40-49 | 15 (22%) | 15 (24%) | 0 (0%) |
| 50-59 | 9 (13%) | 9 (14%) | 0 (0%) |
| 60-69 | 8 (12%) | 8 (13%) | 0 (0%) |
| 70-79 | 5 (7.4%) | 3 (4.8%) | 2 (40%) |
| 80-89 | 3 (4.4%) | 2 (3.2%) | 1 (20%) |
| Biological Sex |  |  |  |
| Male | 46 (68%) | 43 (68%) | 3 (60%) |
| Female | 22 (32%) | 20 (32%) | 2 (40%) |
| Gender |  |  |  |
| Man | 46 (68%) | 43 (68%) | 3 (60%) |
| Woman | 22 (32%) | 20 (32%) | 2 (40%) |
| Place of Residence<sup>2</sup> |  |  |  |
| Northern Manitoba | 65 (96%) | 60 (95%) | 5 (100%) |
| Winnipeg | 1 (1.5%) | 1 (1.6%) | 0 (0%) |
| NR | 2 (2.9%) | 2 (3.2%) | 0 (0%) |
| Received influenza vaccine |  |  |  |
| Yes | 37 (54%) | 33 (52%) | 4 (80%) |
| No | 26 (38%) | 26 (41%) | 0 (0%) |
| Cannot remember | 5 (7.4%) | 4 (6.3%) | 1 (20%) |
| Experienced flu-like symptoms |  |  |  |
| Yes | 30 (44%) | 27 (43%) | 3 (60%) |
| No | 35 (51%) | 33 (52%) | 2 (40%) |
| Cannot remember | 3 (4.4%) | 3 (4.8%) | 0 (0%) |
| Has knowledge of H5N1 |  |  |  |
| Yes | 40 (59%) | 39 (62%) | 1 (20%) |
| No | 24 (35%) | 20 (32%) | 4 (80%) |
| Not sure | 4 (5.9%) | 4 (6.3%) | 0 (0%) |
| Concerned about H5N1 |  |  |  |
| Yes | 13 (19%) | 12 (19%) | 1 (20%) |
| No | 36 (53%) | 34 (54%) | 2 (40%) |
| Not sure | 18 (26%) | 16 (25%) | 2 (40%) |
| NR | 1 (1.5%) | 1 (1.6%) | 0 (0%) |

<sup>1</sup> Median (Q1, Q3); n (%)
<sup>2</sup> NR: not reported

DBS samples were collected by capillary finger prick using a contact-activated lancet, spotted on protein saver cards (Cytiva, 10534612), sealed in a gas-impermeable sachet with desiccant, and stored at −20°C. DBS samples were eluted in phosphate-buffered saline and run on the commercially available Meso Scale Discovery (MSD) Influenza H5 Bridging Serology electrochemiluminescence immunoassay with the use of the H1 Cross-Reactivity Blocker (MSD, R93BL-1) to detect anti-H5 antibodies (MSD, K150AXJU-22) (11). The MSD assay was performed according to manufacturer’s instructions and data were obtained using the MesoScale QuickPlex Q60.

Given that over 50% of samples had calculated concentrations below the assay’s limit of detection, seroreactivity cutoffs were determined using raw signal values to avoid zero-inflating the data. Outliers were determined conservatively using the three-sigma rule, where the mean +/-three standard deviations was calculated using the signal for all samples. Accordingly, samples with a signal above 137.6 were considered outliers (*n*=3) and were excluded from calculations for the seroreactivity cutoff. Calculation of three standard deviations above the mean from the remaining samples (*n*=65) resulted in a seroreactivity cutoff signal value of 57.8.

Data analysis was performed using MSD Discovery Workbench (version 6.0.24) and R Studio (version 2025.05.1). The linear range of the MSD assay was determined using a standard curve, where the first point of the standard curve (H5 Control 1.1 at a 1:10 dilution) was assigned 1000 arbitrary units per millilitre (AU/mL). Linearity of the standard curve was determined by linear regression. According to the linear range of the assay, 40 samples were below the limit of detection (58.8%, *n*=40/68).

Using the signal cutoff value of 57.8, five of the 68 DBS samples (7.4%) were positive for anti-H5 antibodies (**Figure 1**). None of the individuals with H5 antibodies reported exposure to domestic or commercial poultry (**Table 1**). A significant sex bias was present in the study population (χ^2^ = 8.47, *p*-value = 0.0036, Pearson’s Chi-squared test); more individuals identified as male (68%, *n*=46/68) than female (32%, *n*=22/68). Nearly all the participants reported being residents of Manitoba (97%, *n*=66/68) (**Table 1**).

**Figure 1.**
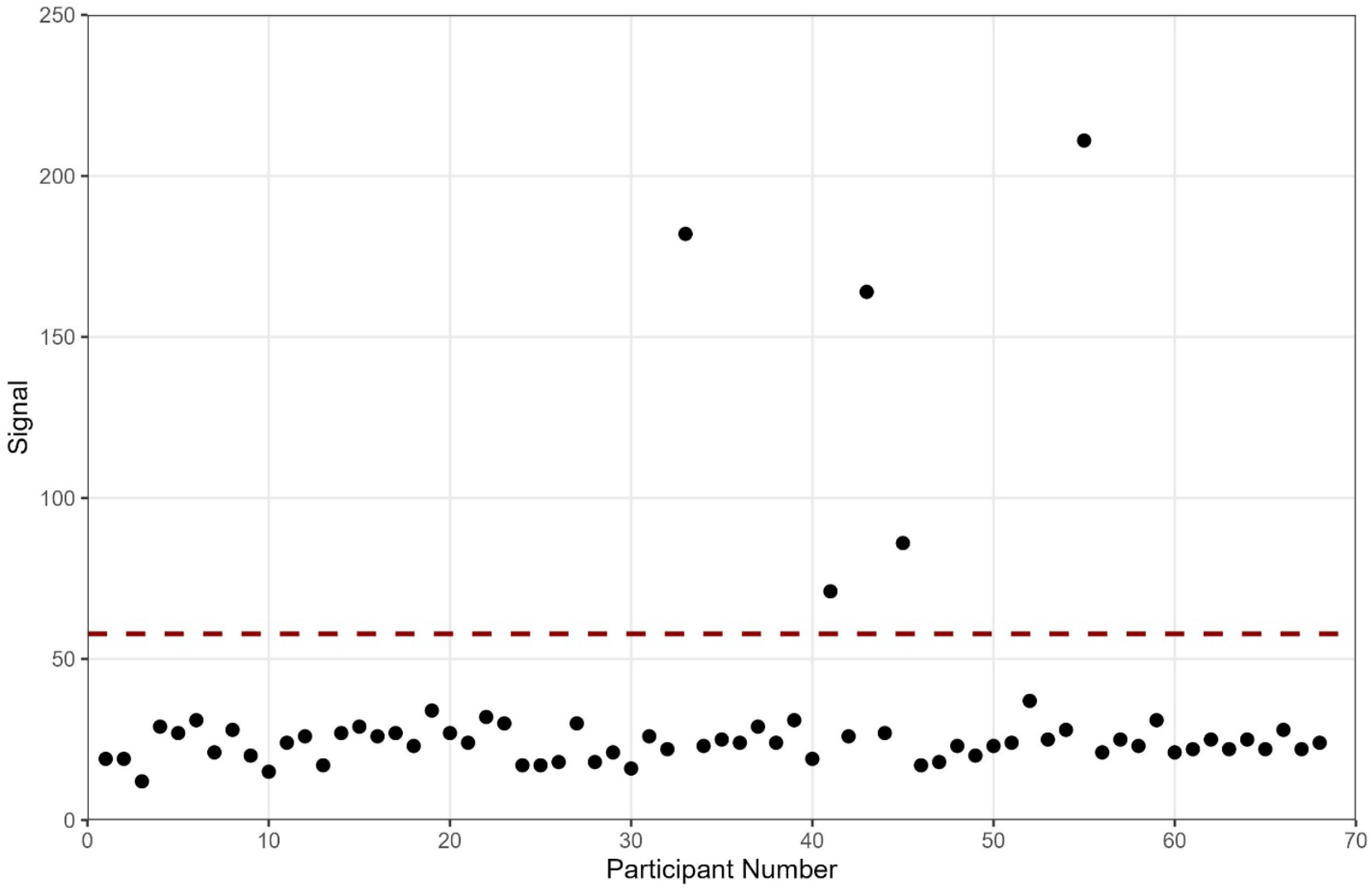
Detection of anti-H5 antibodies from DBS samples. Dotted red line indicates the seroreactivity cutoff signal value of 57.8.

Of the individuals with anti-H5 antibodies (*n*=5), all were involved in the cooking of hunted animals, and most (80%, *n*=4/5) were also involved in hunting/slaughter, cleaning, plucking, and butchering of geese and ungulates (moose, deer, caribou). Ducks were hunted by 3/5 (60%) individuals with anti-H5 antibodies and small game were hunted by 4/5 (80%). These participants all reported regular annual hunting < 2 months per year (100%, *n*=5/5), all of them doing so in the spring season (100%, *n*=5/5) and one reporting hunting in the fall (20%, *n*=1/5) (**Table 2**). Three of the individuals with anti-H5 antibodies reported experiencing flu-like symptoms in the past year (60%, *n*=3/5) (**Table 1**).

**Table 2.** Data from participants regarding hunting and hunting-related activities, frequency, and locations.

| Characteristic | Overall<br>N = 68<sup>1</sup> | Seronegative<br>N = 63<sup>1</sup> | Seropositive<br>N = 5<sup>1</sup> |
| --- | --- | --- | --- |
| Activities |  |  |  |
| Hunting & Slaughter | 45 (66%) | 41 (65%) | 4 (80%) |
| Live Trapping | 15 (22%) | 15 (24%) | 0 (0%) |
| Cleaning | 55 (81%) | 51 (81%) | 4 (80%) |
| Plucking | 55 (81%) | 51 (81%) | 4 (80%) |
| Butchering | 48 (71%) | 44 (70%) | 4 (80%) |
| Collecting Eggs | 24 (35%) | 21 (33%) | 3 (60%) |
| Cooking | 61 (90%) | 56 (89%) | 5 (100%) |
| Domestic Poultry | 8 (12%) | 8 (13%) | 0 (0%) |
| Commercial Poultry | 1 (1.5%) | 1 (1.6%) | 0 (0%) |
| Animal Types<sup>2</sup> |  |  |  |
| Ducks | 27 (40%) | 24 (38%) | 3 (60%) |
| Geese | 67 (99%) | 62 (98%) | 5 (100%) |
| Sea Ducks & Sea Birds | 5 (7.4%) | 4 (6.3%) | 1 (20%) |
| Land Fowl | 22 (32%) | 22 (35%) | 0 (0%) |
| Ungulates | 52 (76%) | 47 (75%) | 5 (100%) |
| Bears | 4 (5.9%) | 4 (6.3%) | 0 (0%) |
| Small Game | 23 (34%) | 19 (30%) | 4 (80%) |
| Other Big Game | 16 (24%) | 15 (24%) | 1 (20%) |
| Marine Mammals | 6 (8.8%) | 6 (9.5%) | 0 (0%) |
| Other | 2 (2.9%) | 2 (3.2%) | 0 (0%) |
| Years of Hunting Experience |  |  |  |
| <2 | 6 (8.8%) | 6 (9.5%) | 0 (0%) |
| 3-5 | 8 (12%) | 7 (11%) | 1 (20%) |
| 6-10 | 9 (13%) | 9 (14%) | 0 (0%) |
| 11-24 | 14 (21%) | 13 (21%) | 1 (20%) |
| 25+ | 30 (44%) | 27 (43%) | 3 (60%) |
| NR | 1 (1.5%) | 1 (1.6%) | 0 (0%) |
| Frequency of Hunting (months per year) |  |  |  |
| <2 | 40 (59%) | 35 (56%) | 5 (100%) |
| 2-5 | 17 (25%) | 17 (27%) | 0 (0%) |
| >5 | 8 (12%) | 8 (13%) | 0 (0%) |
| NR | 3 (4.4%) | 3 (4.8%) | 0 (0%) |
| Hunting Location |  |  |  |
| Northern Manitoba | 65 (96%) | 60 (95%) | 5 (100%) |
| Southern Manitoba | 3 (4.4%) | 3 (4.8%) | 0 (0%) |
| Other Province or Territory | 1 (1.5%) | 1 (1.6%) | 0 (0%) |
| Seasons Participating in Hunting |  |  |  |
| Spring | 63 (93%) | 58 (92%) | 5 (100%) |
| Summer | 16 (24%) | 15 (24%) | 1 (20%) |
| Fall | 37 (54%) | 36 (57%) | 1 (20%) |
| Winter | 26 (38%) | 25 (40%) | 1 (20%) |
| PPE Usage during Handling |  |  |  |
| Always | 14 (21%) | 11 (17%) | 3 (60%) |
| Sometimes | 16 (24%) | 14 (22%) | 2 (40%) |
| No | 36 (53%) | 36 (57%) | 0 (0%) |
| NR | 2 (2.9%) | 2 (3.2%) | 0 (0%) |
| Wash Hands & Change Clothes after Handling |  |  |  |
| Always | 50 (74%) | 46 (73%) | 4 (80%) |
| Sometimes | 13 (19%) | 12 (19%) | 1 (20%) |
| No | 3 (4.4%) | 3 (4.8%) | 0 (0%) |
| NR | 2 (2.9%) | 2 (3.2%) | 0 (0%) |

<sup>1</sup> n (%)
<sup>2</sup> Other includes fish (N=1) and sandhill cranes (N=1).

Importantly, within the overall study population, knowledge of avian influenza was relatively low, with only 59% (*n*=40/68) of participants reporting being aware of H5N1 viruses. Of those who had heard about avian influenza, 32.5% (*n*=13/40) reported being concerned, highlighting the need for additional communication, engagement, and outreach activities, particularly among individuals involved in hunting (**Table 1**).

This seroprevalence study contrasts with a previous human study performed before the introduction of HPAI clade 2.3.4.4b H5N1, which reported no detections of anti-H5 antibodies in the northern United States (12). However, our findings align with more recent studies following the introduction of clade 2.3.4.4b in the United States, reporting up to 14% seroreactivity among individuals who have had close contact with H5N1-susceptible or infected animals (13,14).

The relatively small sample size of the study provides limited power for statistical analyses, and given that the study was conducted in a small region of Manitoba, future larger-scale studies are needed to evaluate how commonly people with exposure to wild birds may be infected by H5N1. Unfortunately, the durability of the antibody response to H5N1 is unknown, therefore we can only conclude that the five seroreactive individuals have been exposed to the virus at some point during their lifetimes, although the durability of antibodies against avian influenza in human populations is unknown. Whether these individuals experienced symptomatic or asymptomatic infections, and whether they would be protected from subsequent re-infection, is also unknown.

## Conclusions

This work indicates that people living and/or hunting in northern Manitoba, Canada have been exposed to H5Nx viruses. Knowledge and concerns about H5N1 among study participants was low, indicating a need for more robust and accessible public health information. Future work including sampling a larger number of individuals and performing longitudinal follow-up to determine how antibody levels change over time would be of significant value. Overall, this work adds to the growing body of literature that aims to understand the true extent of spillovers of avian-origin influenza viruses into humans.

## Data Availability

All data produced in the present study are available upon reasonable request to the authors

## Acknowledgements

We thank the community and individual participants for their support of this work, and Dr. Jordan Wight (Public Health Agency of Canada) for his consultation regarding positivity cutoffs and feedback on the manuscript. We gratefully acknowledge the research assistance of Victoria McEwan and Chris Benson.

## About the Author

Dr. Hannah Wallace is the Lead Research Associate in the Kindrachuk Laboratory at the University of Manitoba. Her research focus is on understanding the pathogenesis of emerging viruses. Ms. Morgan Hiebert is a laboratory technicican in the Kindrachuk Laboratory. Her research interests include viral and bacterial infections among under-served populations.

