## Appendix 1 for "Detection of Anti-H5 Antibodies in People with Exposure to Wild Birds in Northern Canada"

### Assessment of Past Human Exposure to Avian Influenza in Northern Communities

Focused questionnaire to compliment serosurvey using dried blood spots, for individuals who have close interactions with animals that are known to be infected with influenza A viruses. For this questionnaire, specifically people who are involved in hunting, trapping, and/or preparation of hunted/trapped animals in Northern Canada

**General Demographics:**

1. Age:
2. Sex:
3. Gender Identity:
4. Place of residence (town/reserve/nearest town):

**Hunting/Handling :**

5. What types of hunting/trapping-related activities have you participated in in the last 12 months? Check all that apply.
   - ☐ Hunting (slaughter)
   - ☐ Live trapping of animals
   - ☐ Cleaning
   - ☐ Plucking
   - ☐ Butchering
   - ☐ Collecting eggs
   - ☐ Cooking
6. What kinds of animals do you hunt/trap or prepare? Check all that apply.
   - ☐ Ducks (ex. mallards, northern pintails, teals, scaups)
   - ☐ Geese (ex. Canada geese, snow geese)
   - ☐ Sea ducks or seabirds (ex. murres, long-tailed ducks)
   - ☐ Land fowl (ex. grouse, pheasants, wild turkey)
   - ☐ Ungulates (ex. deer, elk, moose)
   - ☐ Bears
   - ☐ Small game (ex. rabbits, squirls, foxes)
   - ☐ Other big game (ex. wolves, coyotes)
   - ☐ Marine mammals (ex. seals, whales)
   - ☐ Other: ________________________________
7. How many years of experience do you have hunting/trapping/handling?
   - ☐ < 2 Years
   - ☐ 3 - 5 Years

☐ 6 - 10 Years
☐ 11 – 24 Years
☐ > 25 Years

8. How frequently do you hunt/handle wild birds?
   ☐ < 2 months per year
   ☐ 2 - 5 months per year
   ☐ > 5 months per year

9. Where do you hunt/handle birds? (check all that apply)
   ☐ Around Churchill
   ☐ around other places in Northern Manitoba
   ☐ around places in Southern Manitoba
   ☐ in other Canadian provinces/territories
   Name which one(s): ______________________________

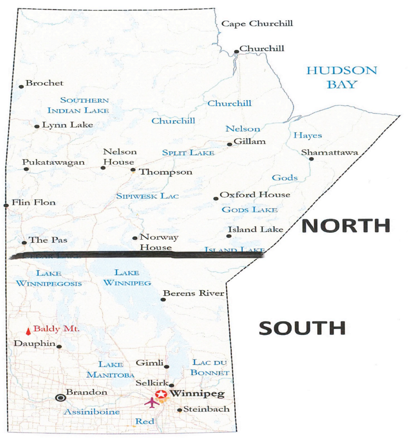


10. What season(s) do you participate in hunting/trapping/handling? Select all that apply.

    ☐ Spring (March, April, May)
    ☐ Summer (June, July, August)
    ☐ Fall (September, October, November)
    ☐ Winter (December, January, February)

11. Do you use any protective equipment when you hunt/trap/handle wild birds or other wild animals? (e.g. gloves, eye covering like goggles, mask, other)

    ☐ Yes, I **always** use:______________________________
    ☐ Yes, I **sometimes** use:______________________________
    ☐ No

12. Do you wash your hands and change your clothes after hunting/trapping/handling game?
    ☐ Yes, Always
    ☐ Yes, Sometimes

☐ No
☐ Prefer not to say

13. Have you ever kept domestic poultry (chickens, ducks, geese) at your home or worked in a commercial poultry plant? (Check all that apply)

☐ Yes – domestic poultry
☐ Yes – commercial poultry
☐ No

**Influenza-specific:**

14. Did you get the influenza vaccine (also known as the flu shot) in the last 12 months?

☐ Yes
☐ No
☐ Cannot remember

15. Have you had flu-like symptoms (e.g. cough, fever, headache, chills, vomiting) at any time in the past 12 months?

☐ Yes
☐ No
☐ Cannot remember

16. Have you heard of highly pathogenic avian influenza (also known as H5N1/Bird Flu/HPAI/AIV)?

☐ Yes
☐ No
☐ Not sure

17. Are you concerned about highly pathogenic avian influenza (also known as H5N1/Bird Flu/HPAI/AIV)?

☐ Yes
☐ No
☐ Not sure
